# Two Decades of Rheumatology Research (2000-2023): A Dynamic Topic Modeling Perspective

**DOI:** 10.1101/2024.06.06.24308533

**Authors:** Alfredo Madrid-García, Dalifer Freites-Núñez, Luis Rodríguez-Rodríguez

## Abstract

**Background:** Rheumatology has experience notably changes in last decades. New drugs, including biologic agents and janus kinase inhibitors, have bloosom. Concepts such as *window of opportunity*, *arthralgia suspicious for progression*, or *difficult-to-treat rheumatoid arthritis* have appeared; and new management approaches and strategies such as *treat-to-target* have become popular. Statistical learning methods, gene therapy, telemedicine or precision medicine are other advancements that have gained relevance in the field. To better characterise the research landscape and advances in rheumatology, automatic and efficient approaches based on natural language processing should be used. The objective of this study is to use topic modeling techniques to uncover key topics and trends in the rheumatology research conducted in the last 23 years.

**Methods:** This study analysed 96,004 abstracts published between 2000 and December 31, 2023, drawn from 34 specialised rheumatology journals obtained from PubMed. BERTopic, a novel topic modeling approach that considers semantic relationships among words and their context, was used to uncover topics. Up to 30 different models were trained. Based on the number of topics, outliers and topic coherence score, two of them were finally selected, and the topics manually labeled by two rheumatologists. Word clouds and hierarchical clustering visualizations were computed. Finally, hot and cold trends were identified using linear regression models.

**Results:** Abstracts were classified into 45 and 47 topics. The most frequent topics were rheumatoid arthritis, systemic lupus erythematosus and osteoarthritis. Expected topics such as COVID-19 or JAK inhibitors were identified after conducting the dynamic topic modeling. Topics such as spinal surgery or bone fractures have gained relevance in last years, however, antiphospholipid syndrome, or septic arthritis have lost momentum.

**Conclusions:** Our study utilized advanced natural language processing techniques to analyse the rheumatology research landscape, and identify key themes and emerging trends. The results highlight the dynamic and varied nature of rheumatology research, illustrating how interest in certain topics have shifted over time.

## 1 Introduction

Over the past decades the volume of academic literature has experienced significant growth Thelwall and Sud [2022], Bornmann et al. [2021]. The field of rheumatic and musculoskeletal diseases (RMDs) has not been immune to this growth, Figure 1. Moreover, RMDs have undergone an unprecedented change in recent years. To begin with, a drug development revolution took place in the early 2000s -which is still active today-, with the arrival of promising drugs such as biologic agents or janus kinase inhibitors Olsen and Stein [2004], Smolen [2020], Kerrigan and McInnes [2020]. Furthermore, the adoption of therapeutic strategies, such as treat-to-target van Vollenhoven [2019], the earlier initiation of disease-modifying treatments, or the paradigm shift in how diseases are analysed, not only by their mortality rate, but also by their disability, propitiated a new scenario for rheumatic and musculoskeletal conditions Kyu et al. [2018], James et al. [2018]. Concepts such as the window of opportunity Burgers et al. [2019], arthralgia suspicious for progression van Steenbergen et al. 2017, erosive disease Van Der Heijde et al. [2013], or difficult-to-treat rheumatoid arthritis Nagy et al. [2021] have gained momentum.

**Figure 1:**
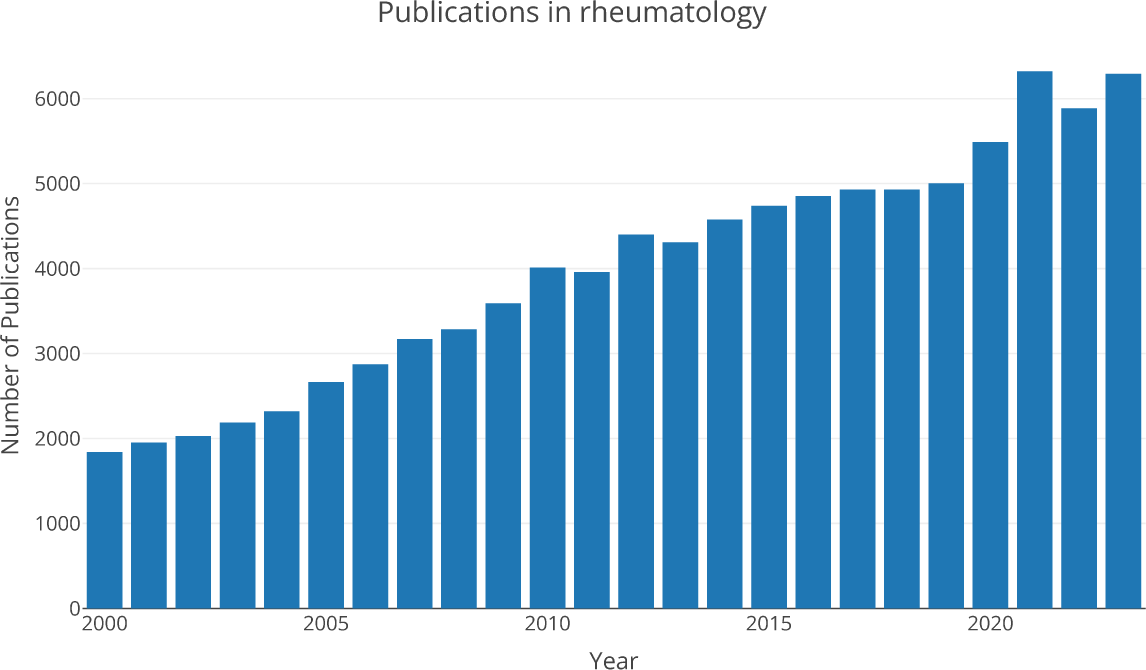
Number of rheumatology-related publications until December 31^st^ 2023.

In this context of continuous change, we hypothesise that the study of trends in scientific publications could be beneficial to better understand the historical research priorities in rheumatology and the evolving landscape of RMDs management and treatment. However, with almost 100,000 original articles published in the last 23 years, the process of comprehending and identifying the main trends is becoming increasingly challenging.

Conventional review methods can be labor-intensive, overwhelming or unfeasible, and non-exhaustive. Hence, we propose the use of modern natural language processing techniques to characterise the evolution of the research topics addressed over time in rheumatology scientific publications. Topic modeling (TM) techniques, are ideally suited for this, as they can model the evolution of topics over time. Briefly, TM is a suite of unsupervised learning algorithms (i.e., no tags/labels are provided with the input data), within the field of machine learning, designed to identify prevalent topics within a corpus of documents, usually through probabilistic methods Churchill and Singh [2022], Abdelrazek et al. [2023]. In that collection, the documents are observed while the topic structure (i.e., the topics, per-document topic distributions, and the per-document per-word topic assignments) is hidden structure Blei [2011, 2012]. The outcome of a typical topic modeling algorithm is clusters of related words. These techniques operate under the assumption that each topic is defined by a distinct collection of words, and that a document consists of a blend of multiple topics in varying proportions. One of the most widely used TM techniques is Latent Dirichlet Allocation (LDA), a generative probabilistic model. However, with the recent advances in NLP and the introduction of the transformer’s architecture, new TM techniques that consider semantic relationships among words and their context have arouse (i.e. BERTopic).

Consequently, this research study seeks to address the question: *How has rheumatology research evolved in recent years?* To do so, we employ BERTopic to uncover trends related to rheumatology and to explore the publication landscape of rheumatology research within the scientific literature over the past two decades.

## 2 State of the art

TM has been used in a multitude of fields, including social networks, software engineering, crime science, political science, geography, medicine, and linguistics Jelodar et al. [2019]. Additionally, it has proven effective in analyzing historical documents such as newspapers and humanistic texts Boyd-Graber et al. [2017], as well as in educational research Mulunda et al. [2018], and the study of organizational phenomena Valeri [2021].

TM has been widely applied in rheumatology research.

The authors in Tedeschi et al. [2021] employed a TM approach, sureLDA, followed by penalised regression, to predict pseudogout probability in large datasets. TM was also applied to characterise the temporal evolution of ANCA-associated Vasculitis (AAV) in Wang et al. [2021]. Temporal trends, in more than 113,000 clinical notes, before and after the treatment initiation date for a diagnosis of AAV, were modelled with LDA, finding 90 different topics that included diagnosis (e.g., granulomatosis with polyangiitis), treatments (e.g., AAV specific-treatment), and comorbidities and complications of AAV (e.g., glomerulonephritis, infections, skin lesions).

A prior study, Dzubur et al. [2019], explored the application of TM to understand the concerns and perceptions of patients with ankylosing spondylitis regarding biologic therapies. The researchers analyzed over 25,000 social media posts using LDA and identified 112 topics. Medication uncertainty, lack of trust in physician’s decisions, patient worries and seeking alternative treatments highlighted were those most prevalent.

On its behalf, in Li and Yacyshyn [2023], scholars analysed the posts published over the course of a year in the Reddit subforum ‘r/Behcet’ to investigate the perspectives and experiences of people affected by Behcet’s disease. The authors identified 6 themes and 16 subthemes, including *finding connectedness through shared experiences, the struggles of the diagnostic odyssey and sharing or inquiring about symptoms*.

In noa [2023a], the authors pursue to uncover the themes present in the Electronic Health Record (EHR) of patients with rheumatoid arthritis (RA) prior to the start of targeted treatments, and to explore their relationship with the subsequent course of treatment. On the other hand, in noa [2023b] the authors evaluated two social media communities, a Facebook group, and a public subreddit, ‘r/gout’, identified 30 topics and conduct sentiment analysis.

Moreover, investigators in noa 2018 characterised systemic lupus erythematosus (SLE) patients’ experiencies in an online health community by applying LDA in free text data extracted from *PatientsLikeMe* community.

Eventually, in Sperl et al. 2022, LDA was applied to analyze responses to open-ended questions from an online survey designed to assess motivations among health professionals for participating in post-graduate rheumatology education; and to identify barriers and facilitators for participation in current EULAR educational offerings.

Table 1 shows the most relevant characteristics of each study discussed above.

**Table 1:** Rheumatology studies in which topic modeling has been employed. EHR: Electronic Health Record. LDA: Latent Dirichlet Allocation.

| Author | Disease | Data source | Number of data points / patients | Topics | Algorithm |
| --- | --- | --- | --- | --- | --- |
| Sara K Tedeschi et al. (2021) Tedeschi et al. [2021] | Pseudogout | EHR | 30,089 patients | - | sureLDA |
| Liqin Wang et al. (2021) Wang et al. [2021] | Vasculitis | EHR | 113,000 notes | 90 | LDA |
| Eldin Dzubur et al. (2019) Dzubur et al. [2019] | Ankylosing spondylitis | Social media data | 27,416 posts from 601 social media sites | 112 | LDA |
| Jenny Xiaoyu Li et al. Li and Yacyshyn [2023] (2023) | Behçet | Social media data (Reddit) | 196 threads | 6 | - |
| Jason Tang et al. (2023) noa [2023a] | Rheumatoid arthritis | EHR | 1,102 patients | 4 | LDA |
| Maurice Flurie et al. (2023) noa [2023b] | Gout | Social media data (Facebook and Reddit) | 175,000 posts comments | 30 | - |
| Stephanie Eaneff et al. (2023) Eaneff et al. [2018] | Systemic lupus erythematosus | Online community | 138,409 posts | 150 | LDA |
| L. Sperl et al. (2022) Sperl et al. [2022] | - | Online survey answers | 667 answers | - | - |

## 3 Materials and Methods

### 3.1 Materials

Data from the *RheumaLpack* corpus Madrid et al. 2024, which includes 96,004 rheumatology-related abstracts along with associated metadata, up to 19 variables including *title, PMID/DOI, abstract, publication year, journal, keywords, or volume*, were extracted. These abstracts were compiled from original articles indexed in PubMed from January 1, 2000, to December 31, 2023; and came from 34 rheumatology-specific journals, as identified by the Journal Citation Reports (JCR), see Supplementary Tables 1 and 2. R’s *rentrez* library was used to collect the data.

BERTopic was used for topic modeling Grootendorst [2022]. This technique generates topic representations through three steps:

- **Document embeddings**: Unlike LDA, a probabilistic topic modeling approach, BERTopic utilizes pre-trained language models to create representations that can be compared semantically. Therefore, clusters of semantically similar documents, abstracts, are created.
- **Document clustering**: to overcome the *curse of dimensionality*, the dimensionality of document embeddings generated in previous step is reduced. Uniform Manifold Approximation and Projection (UMAP) algorithm is commonly employed for that purpose. After that, the reduced embeddings are clustered using HDBSCAN algorithm. This is a soft-clustering approach that prevent the merging of dissimilar topics, this is, the algorithm strategically generates outliers (i.e., documents that do not fall within any of the created topics) to handle the noise. In BERTopic, these outliers are tagged as *topic “-1”*.
- **Topic representation**: each cluster is assigned to a topic. To measure the relevance of each term (i.e., word) in a topic, the class based TF-IDF (c-TF-IDF) approach was used. This is a modification of TF-IDF, that models the importance of words in clusters instead of in documents. With c-TF-IDF, it is also possible to model how topics evolved over time following a *dynamic topic modeling* approach.

### 3.2 Methodology

The *abstract, title, publication year*, and *journal* information for the 96,004 original articles was retrieved from the *RheumaLpack* corpus. The number of tokens per abstract was computed to guide the selection of the embedding model. This is crucial because texts that exceed the model’s maximum length limit are truncated during the embedding process, leading to a loss of information. Depending on the median token size, two options were considered a) to concatenate the title and the abstract, so only a complete and single text for each article is studied; b) to focus the study solely on abstract information.

Data pre-processing was omitted to preserve the original text structure, which is relevant for transformer-based models to effectively comprehend the context. Hence, stopwords were not omitted. From here onwards, the modular approach of BERTopic was applied, with considerations made for each step:

- Embeddings were calculated to feed the BERTopic model. Two models were considered:

**–** *all-mpnet-base-v2*: sentence-transformer model that maps sentences and paragraphs to a 768 dimensional dense vector space. This model was trained on a 1B sentence pair, comes from the pre-trained MPnet model Song et al. [2020], and was fine-tuned using a contrastive objective. It was the best positioned model in the sentence transformers rank by March 2024 sbe 2024. By default, input text longer than 384 word pieces is truncated. This model has been applied in Ramamoorthy et al. [2024], Ng et al. [2023], Guizzardi et al. [2023], Meaney et al. [2022].

**–** *S-PubMedBert-MS-MARCO*: a sentence-transformer model specially optimised for medical texts Deka et al. [2022]. This model max sequence length is 350. Input text longer than this size is truncated. This embedding model has been used in the past for similar tasks Karabacak and Margetis [2023], Karabacak et al. [2024a,b,c], Ozkara et al. [2023, 2024].

- The embeddings were resized using the UMAP dimensionality reduction algorithm. The algorithm’s parameters were set to default, except for the random_state parameter, which controls the algorithm’s stochastic behavior by fixing a seed. To assess the consistency of the generated topics, three different seeds (i.e., *random_state*) were applied to each tested model. This approach facilitated a comparison across various initializations, using stability as an intrinsic evaluation metric for evaluating performance.
- HDBSCAN was used as the default clustering algorithm. The cluster minimum size (i.e., *min_cluster_size*) was set to 50, 100, 150, 200 and 250 (i.e., minimum number of documents per topic). As this number increases, the number of microclusters decreases, resulting in fewer topics.
- The default vectorizer model, *CountVectorizer*, was chosen to preprocess the topic representations after the documents were assigned to topics. Stopwords and infrequent words were removed in this step. The n-gram range considered was 1-2, meaning that topic representations made up of one or two words were allowed. Other representations were also explored such as *KeyBERTInspired*, *MaximalMarginalRelevance* (i.e., pursues to maximize the diversity of keywords) and *PartOfSpeech* (i.e., extract keywords based on their Part-of-Speech).

The number of words extracted per topic was set to 20 (i.e., *top_n_words*), as the optimal number of words in a topic is between 10 and 20. Beyond this range, topics tend to lose coherence. We explored all potential combinations involving two embedding models (i.e., *all-mpnet-base-v2* and *S-PubMedBert-MS-MARCO*), three different UMAP inicialization states (i.e., seeds 42, 52, and 62), and five cluster minimum size values (i.e., 50, 100, 150, 200 and 250). A total of 2 *∗* 3 *∗* 5 = 30 models were trained.

Two final models were selected for further analysis: one using *all-mpnet-base-v2* and the other using *S-PubMedBert-MS-MARCO*. This selection was based on several criteria, including the number of outliers, the number of topics, and the topic coherence score (i.e., u_mass). The chosen models were required to contain fewer than one third of the total documents classified as outliers (n *<* 32,000), support more than 40 topics, and minimise the u_mass score. This score, is an intrinsic evaluation method (i.e., measures the quality of the topic model itself without considering any specific external task) that evaluates the quality of a topic based on co-occurrences of word pairs Rosner et al. [2014], which was introduced in Mimno et al. [2011]. Other coherence measures were calculated (i.e., c_v, c_nmpi and c_uci) but the final decision was guide by c_umass. Afterwards, outliers were excluded from the analyses.

After analyzing the keywords and the different topics representations, the topics were labelled through a mutual agreement among D.F.N and L.R.R authors. Word clouds were generated to show the keywords linked to the topics and the topics’ distribution. The size of each word is proportional to its relevance in the topic. Hierarchical clustering representations were generated to show how topic embeddings can be combined at various cosine distances. Dynamic topic modeling was employed to explore the evolution of topics over time, using the two selected models.

Eventually, we applied the same methodology described in Karabacak and Margetis [2023] to model trends. The publication year, and the topics probabilities (i.e., the probability of an abstract being classified under a particular topic based on its content) were retrieved. The mean topic probability per publication year and per topic was computed. Bivariate linear regression models were developed for each topic, with the mean topic probability serving as the dependent variable, and the publication year as the independent variable. By examining the slopes of these regression lines, topics were categorized as hot if they had positive slopes and cold if they had negative slopes.

All models were trained in Google Colab, with a T4 GPU and a high-RAM runtime, using Python.

## 4 Results

The median number of tokens per abstract was 375 (Q1: 287, Q3: 442). When combining both abstract and title, the median was 401 (Q1: 310, Q3: 471), therefore, we chose to analyze only the abstract. The number of topics identified by the models ranged from 42 to 296, while the number of initial outliers ranged from 19,075 to 35,332. In Supplementary Table 3, the results of the 30 trained models are shown, including the minimum cluster size, the seed, the number of topics and outliers, and the coherence score values. As the number of topics decreases (and the number of the minimum cluster size increases), the topic coherence scores are better. In *Supplementary Excel File Models Output* the topic number, the count, the default topic name, the different topic representations and the three abstracts that best encapsulate the thematic content of each topic are shown. *Supplementary Excel File Top 5 Topics* shows the five topics with the highest number of documents for all models.

The model that exhibited the lowest u_mass coherence score utilized a minimum cluster size of 250, with seed values of 52 for the *all-mpnet-base-v2* model (−0.279) and 42 for the *S-PubMedBert-MS-MARCO* model (−0.288). A total of 73,736 and 69,316 abstracts were classified into 47 topics and 45 topics for the *all-mpnet-base-v2* and the *S-PubMedBert-MS-MARCO* models, respectively. The remaining documents were classified as outliers and discarded. Tables 2 and 3 present a detailed overview of the topics, outlined by a unique set of keywords that capture their essential themes.

**Table 2:**
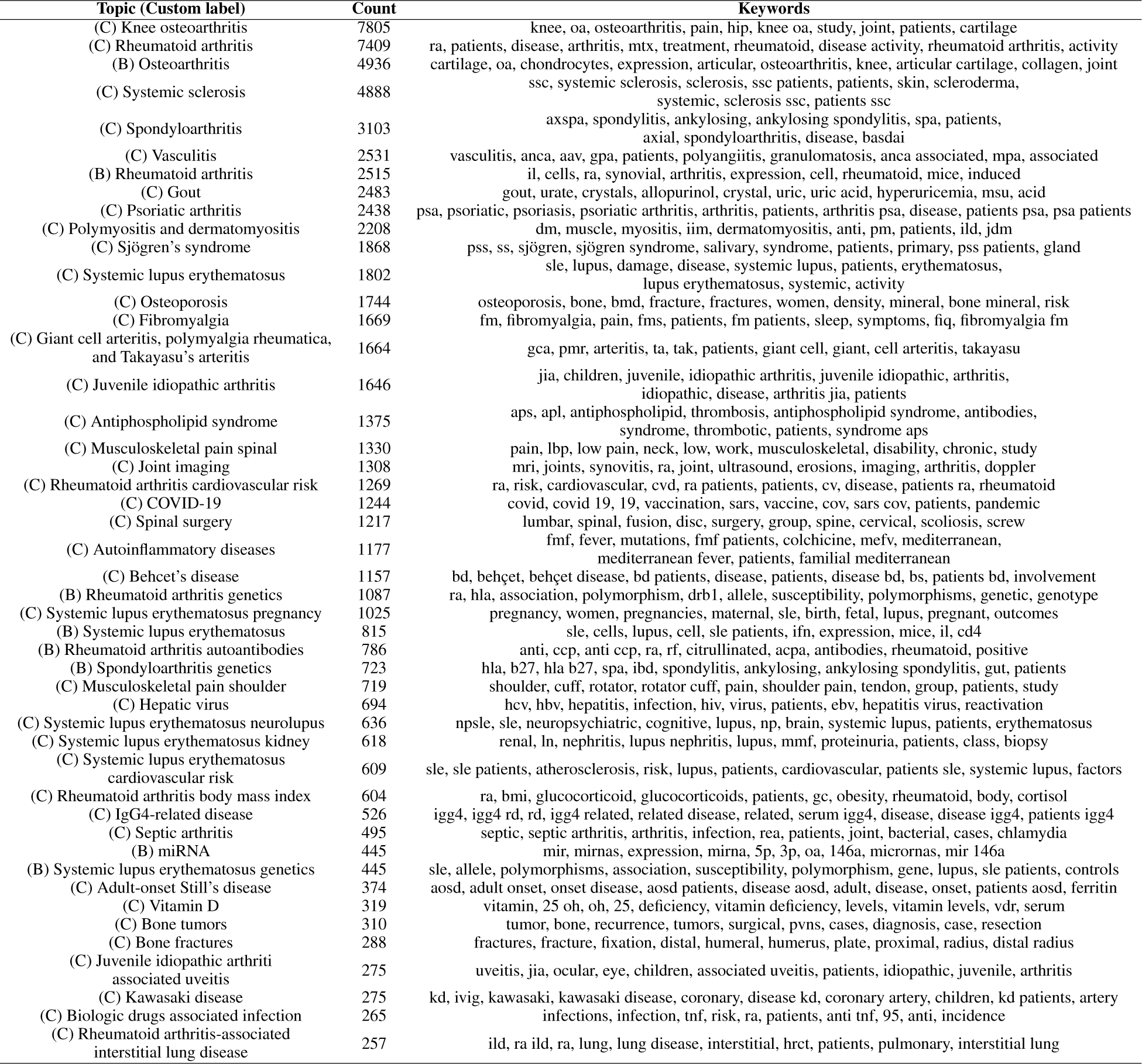
Summary of the topics for the *all-mpnet-base-v2* model.

**Table 3:**
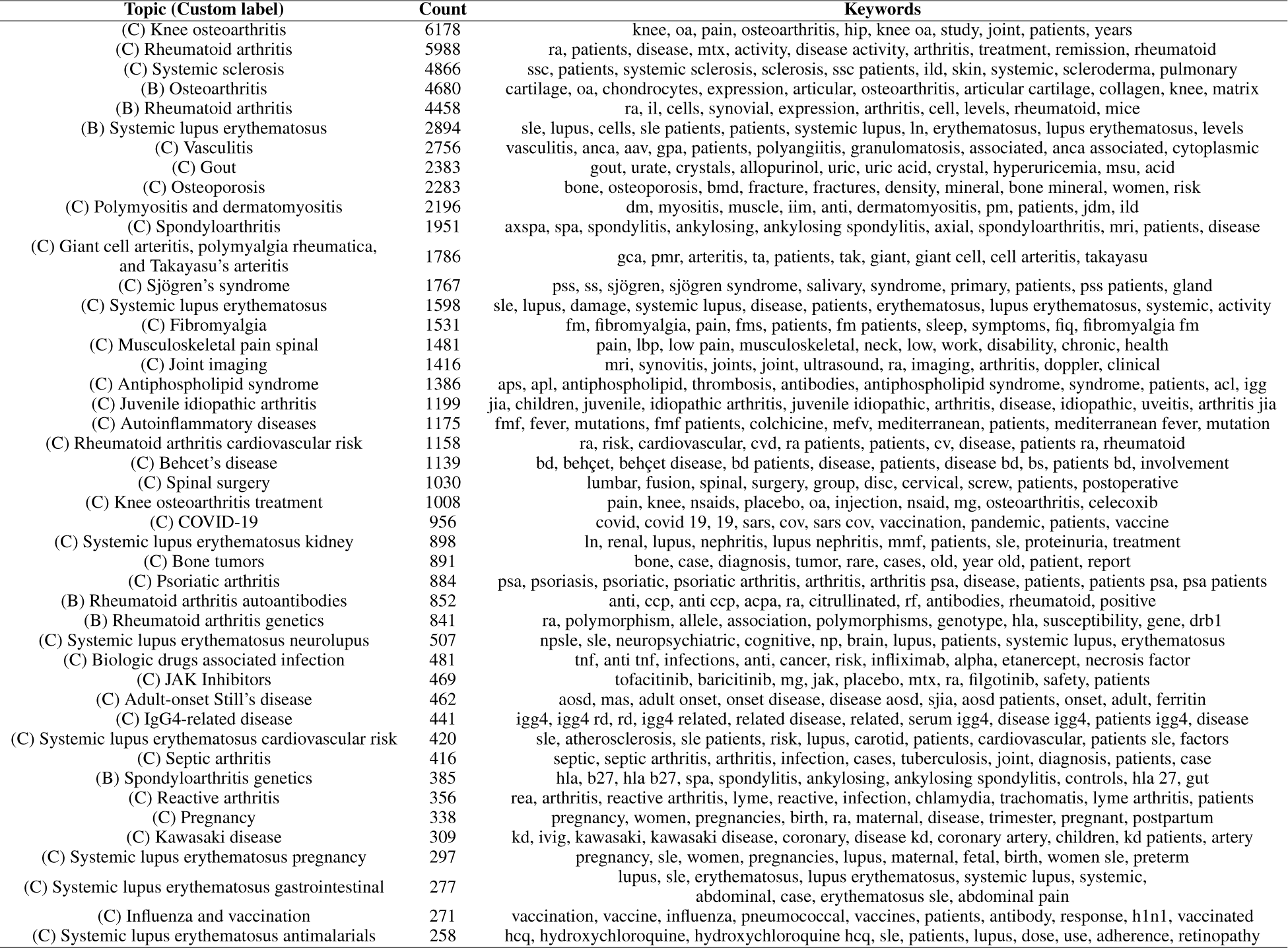
Summary of the topics for the *S-PubMedBert-MS-MARCO* model.

Hierarchical clustering plots and word clouds for the top ten topics are shown in the Supplementary Figures 1 and 2, and 3 and 4, respectively.

Regarding the dynamic modeling of topics, for each model we studied the themes in batches of 10. Figures 2 and 3 show the results. Moreover, a bar chart of the hot and cold topics for the two models is displayed in Figures 4 and 5. Finally a comparison of the topics of the two final models is presented in Supplementary Table 4.

**Figure 2:**
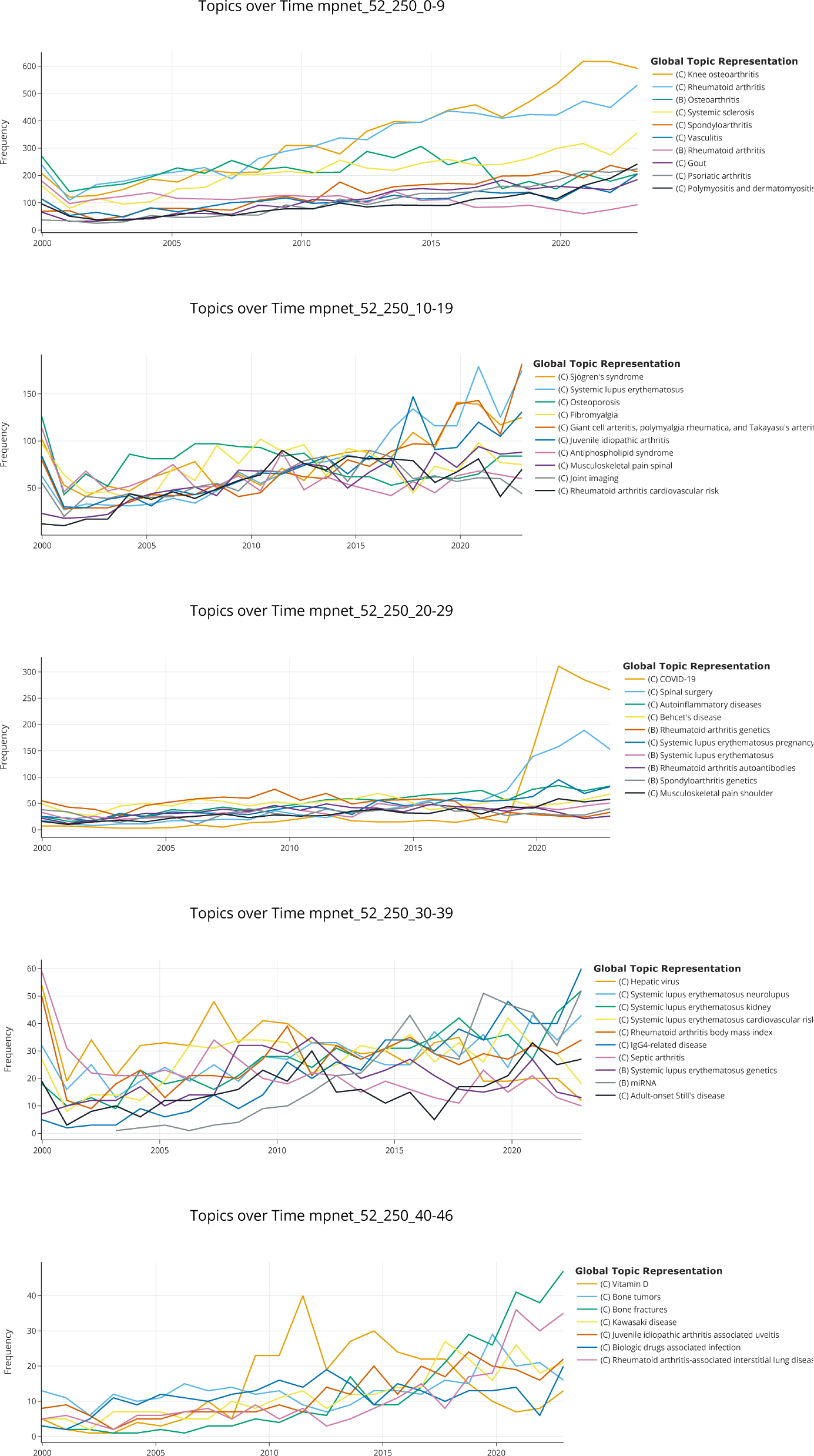
Dynamic topic modeling of the best *all-mpnet-base-v2* model.

**Figure 3:**
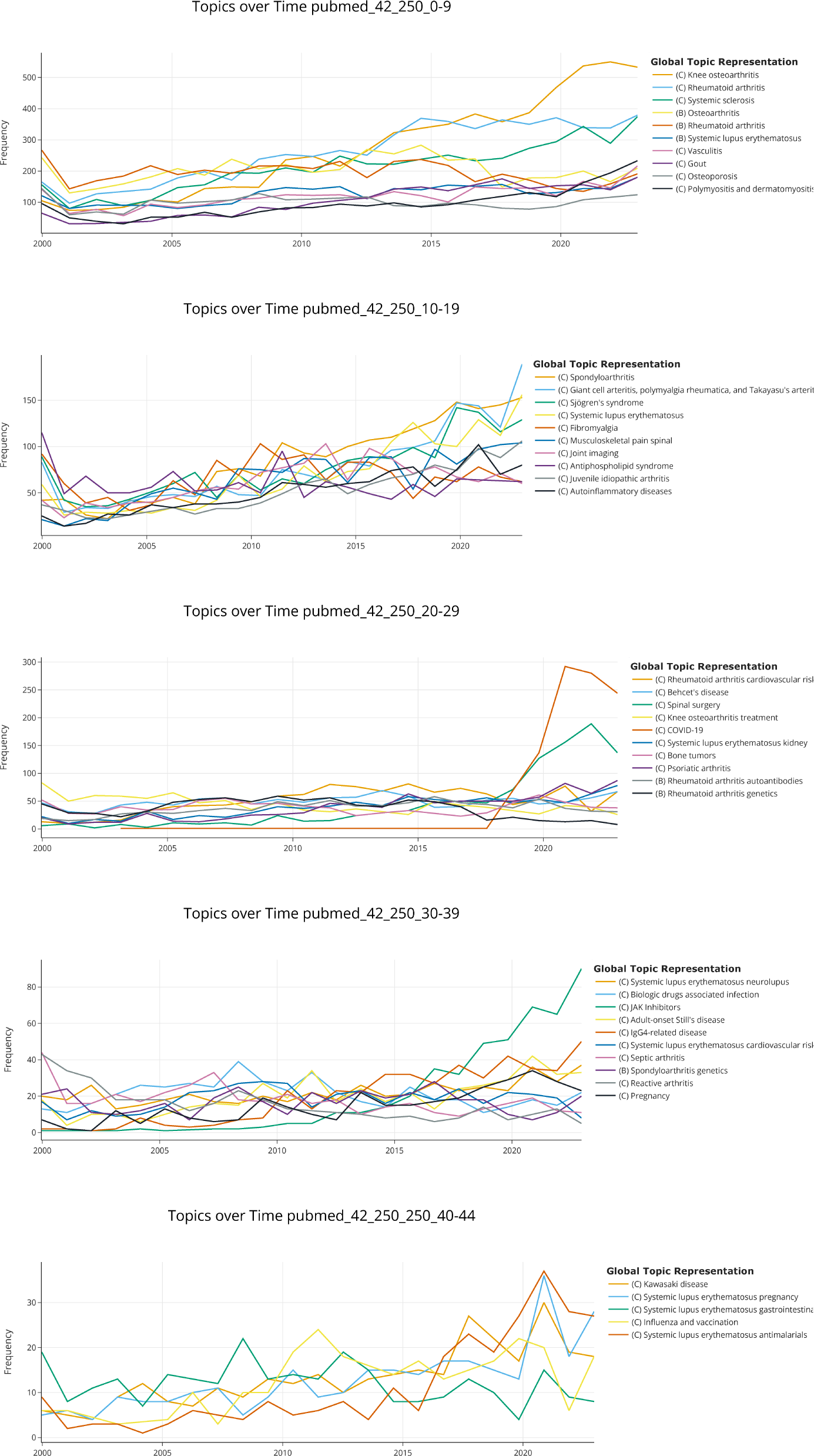
Dynamic topic modeling of the best *S-PubMedBert-MS-MARCO* model.

**Figure 4:**
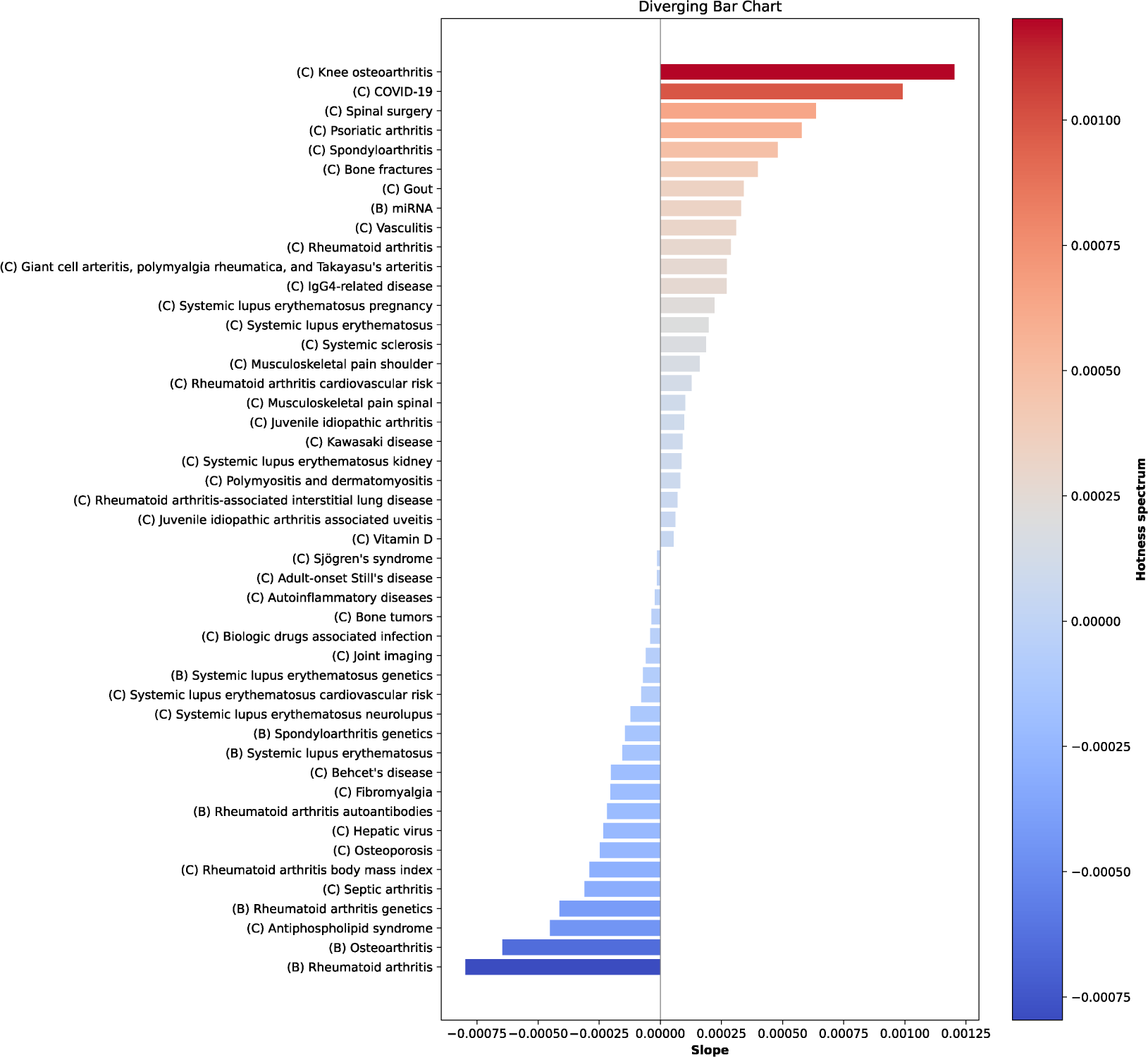
Bar chart of hot and cold topics. *all-mpnet-base-v2* model.

**Figure 5:**
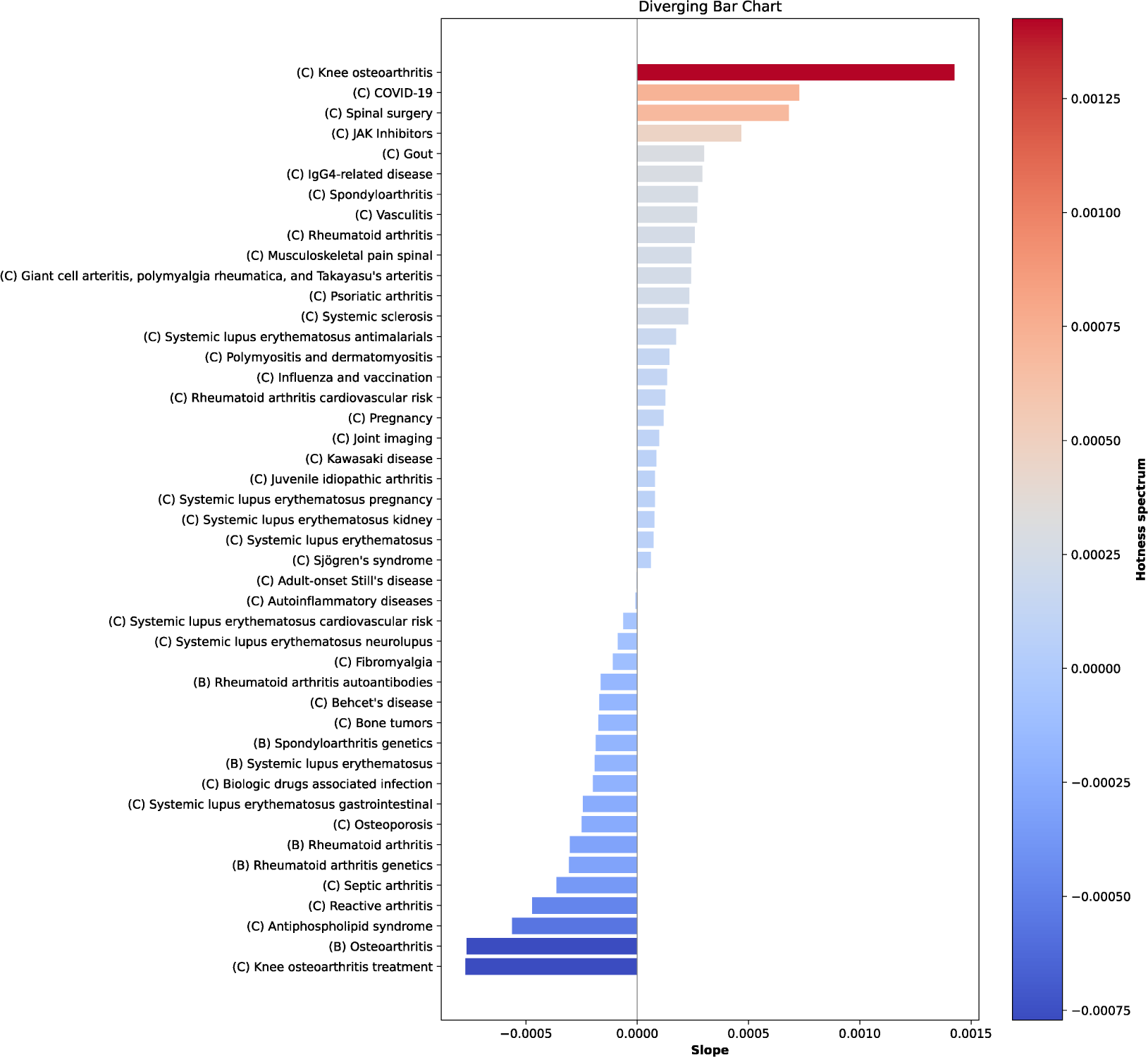
Bar chart of hot and cold topics. *S-PubMedBert-MS-MARCO* model.

## 5 Discussion

### 5.1 Trends in rheumatology research

When comparing the top ten topics identified in the two models, *all-mpnet-base-v2* and *S-PubMedBert-MS-MARCO*, there is considerable overlap between them. This overlap could lend credibility to the findings. For instance, eight of the ten primary topics were consistent across the models, with (C) Knee osteoarthritis, and (C) Rheumatoid arthritis being the most studied topics. The relevance of (C) Spondyloarthritis, (C) Psoriatic arthritis, (B) Systemic lupus erythematosus, and (C) Osteoporosis topics differ between both models. However, when combining all the topics related to RA and SLE, the number of documents is 13,927 and 5,950 for the *all-mpnet-base-v2* model, and 13,297 and 7,149 for the *S-PubMedBert-MS-MARCO*. Therefore, globally, the three most studied topics are: RA, SLE, and osteoarthritis.

Some of the topics expected to be found (e.g., (C) COVID-19 and (C) JAK inhibitors) were present after applying dynamic topic modeling, which further strengthens the reliability of the results. Conversely, other unexpected topics such as (C) Spinal surgery or (C) Bone fractures have gained relevance in recent years. As shown in Figures 5 and 4; (C) Gout, (C) Spondyloarthritis and (C) Psoaritic arthritis are nowadays *hot topics*, whereas (C) Antiphospholipid syndrome, (C) Septic arthritis or (C) Reactive arthritis are *cold topics*.

As the final number of topics is relatively low, no specific topics related to artificial intelligence or new statistical learning techniques that became popular a few years ago, such as trajectory analysis, were identified. However, when analysing models with a higher number of topics such as *all-mpnet-base-v2* (minimum number of cluster: 50, seed: 42) we found the following topics: [learning, machine, algorithms, machine learning, algorithm, ai, deep learning, artificial intelligence, artificial, intelligence]. Something similar occurs with social media data topic: [websites, internet, information, social media, readability, search, media, social, google, online], with telemedicine [app, apps, mobile, smartphone, digital, application, care, health, mhealth, patient], and with wearables: [app, apps, mobile, smartphone, digital, application, care, health, mhealth, patient]. Hence, the use of models with a larger number of topics could be useful to identify new emerging trends. See *Supplementary Excel File Models Output*.

### 5.2 Topic modeling in PubMed abstracts

The use of TM techniques on PubMed abstracts is not new. These methods have been used in different medical fields for trend analysis and for uncovering hidden topics over the past few years. For example, the authors in Sperandeo et al. [2020] evaluated the usage of “personality” and “mental health” terms within the titles and abstracts of articles published in PubMed from 2012 to 2017. The researchers employed LDA on more than 7,500 abstracts and found 30 topics organised in eight hierarchical clusters, concluding that personality is linked to a broad spectrum of conditions. The suitable number of clusters was determined using a 5-fold cross-validation approach.

The authors in Tighe et al. [2020] applied TM on a corpus of more than 200,000 abstracts related to pain. The abstracts collected, retrieved through searches using “pain” [MeSH] term, corresponded to articles published between 1949 and 2017. On this occasion, both LDA and latent semantic indexing techniques were employed. After following a topic coherence strategy, the researchers identified an optimal topic count of 40. One of the conclusions of this research was that TM can be helpful in identifying critical research avenues by evaluating the gaps in the literature concerning a specific topic.

On their behalf, researchers in Abba et al. [2022], focused on the use of TM techniques to uncover hidden topics from 100 years of peer-reviewed hypertension publications (i.e., 1900-2018). LDA was applied to more than 580,000 abstracts. Most of the identified topics, n = 20, fell into four distinct categories: preclinical, epidemiology, complications, and treatment-related studies. Topic trends were evaluated by calculating the annual proportion of abstracts for each topic relative to the cumulative total of articles associated with that topic.

Researchers in Shi et al. [2023] examined artificial intelligence (AI)-related studies published in PubMed, from 2000 to 2022, to highlight the current situation of medical AI research and to provide insights into its future developments. With that aim, scholars downloaded metadata from 307,000 articles, (e.g., title, abstract, journals, authors) and applied LDA to titles and abstracts. They divided the data into intervals of five years, performing unique TM for each period. The authors presented the five main topics in eight different domains of AI. These domains were described by the European Commission Joint Research Centre.

Depression, anxiety, and burnout in academia were studied through BERTopic in Lezhnina [2023]. The authors extracted 2,846 abstracts from PubMed ranging from 1975 to 2023 using a complex query that did not include MeSH terms. Afterwards, the authors compared BERTopic models with different sets of parameters, each of them being run three times. The best model was chosen based on different criteria (i.e., proportion of outliers, topic interpretability, topic coherence, and diversity); this model comprised 27 topics. After studying their evolution, the authors showed, among others, how the COVID-19 pandemic influenced the burnout of medical professionals.

Eventually, in Grubbs et al. [2023] the researchers studied the topics present in a specific academic journal-Gynecologic Oncology-over a thirty-year period (i.e., 1990-2020), as well, as the interest in them over time. With that aim, they used LDA on 11,200 abstracts and determined the number of topics using the coherence score. The best model contained 26 topics, and three of them were merged after manual assessment by three reviewers. Thanks to the experiments carried out, researchers could hypothesise the evolution of some topics related to oncology gynecology for the next years, such as an increase in surgical topics and in epidemiological and health outcomes research topics; and a decrease in chemotherapy and radiation.

As can be seen from the above studies, there is a real interest in uncovering latent topics in medical documentation. In this study, we have demonstrated how dynamic topic modeling can be applied to abstracts indexed in PubMed, and published in Rheumatology journals from 2000 to 2023.

To the best of our knowledge, the BERTopic approach has not been previously applied to examine trends within this medical field. A potentially more intriguing application of dynamic topic modeling would involve its use with EHR data, to characterize the natural history of diseases. This approach was taken a few years ago, but applying LDA over AAV histories Wang et al. [2021].

Furthermore, each clinical note could be categorized into a specific topics. Should there be a requirement for a manual review of the record contents, pre-classifying them by topic could assist physicians in assembling patient cohorts for targeted studies.

Finally, these models could be used as recommendation systems to direct unpublished scientific articles to the journal that maximises their likelihood of publication based on the latent topics contained in the abstract and other structured data (e.g., year, affiliation of the first author).

### 5.3 Limitations

- Biologic agents were introduced in the market in 1999. As our study window begins in 2000 we missed the evolution of this topic, from early experiments and clinical trials to their commercial release.
- Topic modeling involves a degree of subjectivity. The results we showcased suggest that topic modeling can be used to discover and understand research trends, rather than assessing the performance of BERTopic as a topic model.
- BERTopic has some noteworthy limitations, as documented by Grootendorst in 2022 Grootendorst [2022]. One significant limitation is the assumption that each document pertains to only one topic, which overlooks the likelihood of documents covering multiple topics.
- Analyzing multiple journals might offer a more comprehensive view, but it also brings variability from the distinct scopes and editorial standards of each journal. This variability may complicate the analysis of research topics and trends. However, both methods—analyzing a single journal Ozkara et al. [2024] and examining multiple journals Karabacak and Margetis 2023— have been utilized in previous research.
- We have not associated the research trends with other indicators, such as the number of patented products or the volume of clinical trials.

## 6 Conclusion

To our knowledge, this is the first study that uses BERTopic, and dynamic topic modeling to identify the key topics in rheumatology research using a set of abstracts extracted from PubMed. The two sentence embeddings models employed, provided similar results, highlighting the dynamic and varied nature of rheumatology research and illustrating how interest in certain topics has shifted over time. As the number of scientific publications increases, the use of natural language processing techniques will be necessary to efficiently analyze and synthesize information, helping to identify trends, gaps, and emerging areas of interest across various medical fields.

## Data availability statement

All data used in this manuscript is available online at https://pubmed.ncbi.nlm.nih.gov/. Data processing is described in Madrid-García, A., Merino-Barbancho, B., Freites-Núñez, D., Rodríguez-Rodríguez, L., Menasalvas-Ruíz, E., Rodríguez-González, A., & Peñas, A. (2024). From Web to RheumaLpack: Creating a Linguistic Corpus for Exploitation and Knowledge Discovery in Rheumatology. medRxiv, 2024-04.Madrid et al. 2024.

Further inquiries can be directed to the corresponding author.

## CRediT author statement

**Alfredo Madrid-García**: Conceptualization of this study, methodology, coding, review, writing (original draft preparation). **Dalifer Freites Núñez**: Methodology **Luis Rodríguez Rodríguez**: Methodology, review

## Supplementary material files

- Supplementary Excel File Models Output: topics identified in the 30 models trained
- Supplementary Excel File Top 5 Predominant Topics: predominant topics in the 30 models trained

## Supporting information

Supplementary Excel File Top 5 Predominant Topics.xlsx

Supplementary Excel File Models Output.xlsx

## Acknowledgement

The authors would like to thank: Inés Pérez San Cristobal, Anselmo Peñas, and Alejandro Rodríguez González

---

**Supplementary Figure 1:**
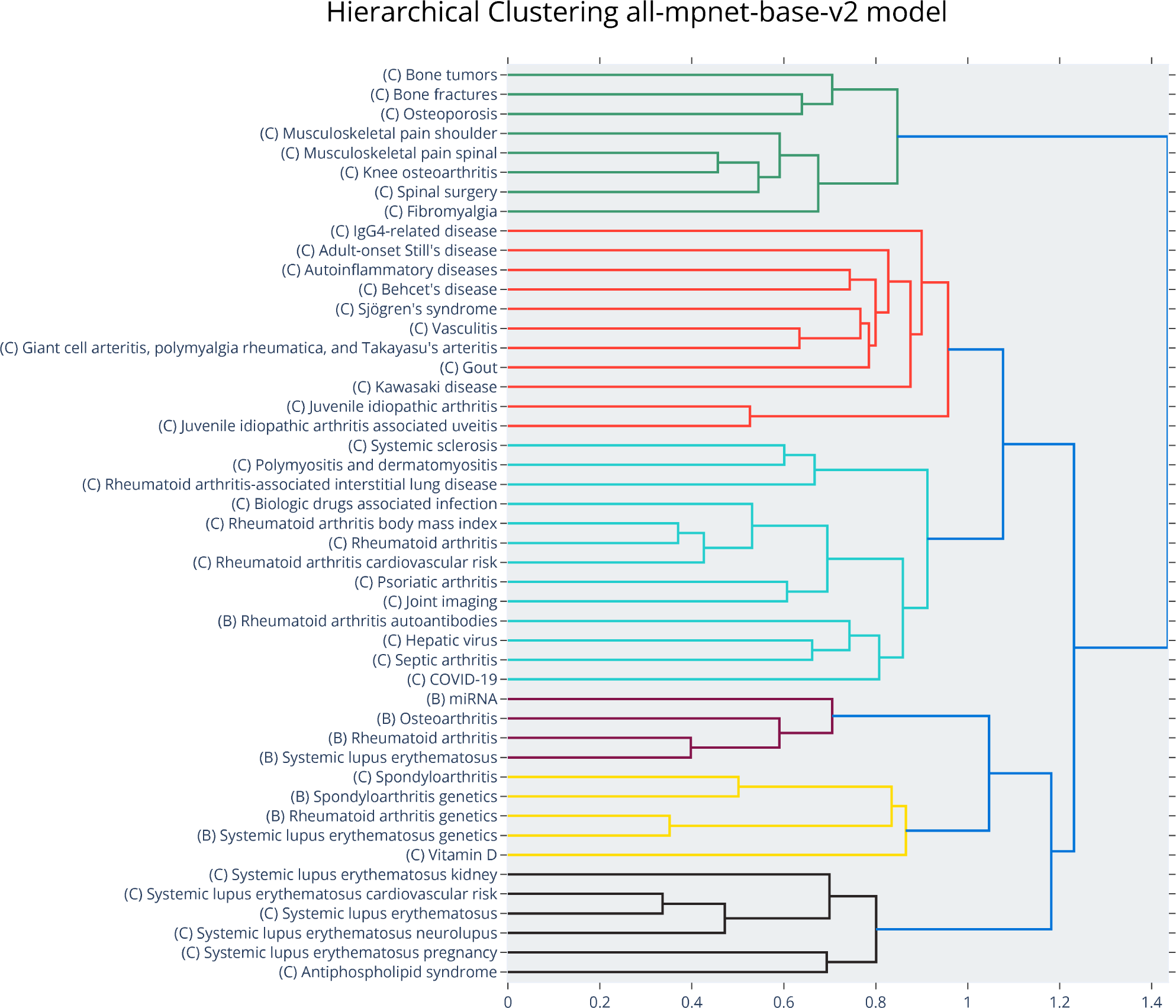
Hierarchical structure of the topics labeled with the agreed label. Best *all-mpnet-base-v2 model*.

**Supplementary Figure 2:**
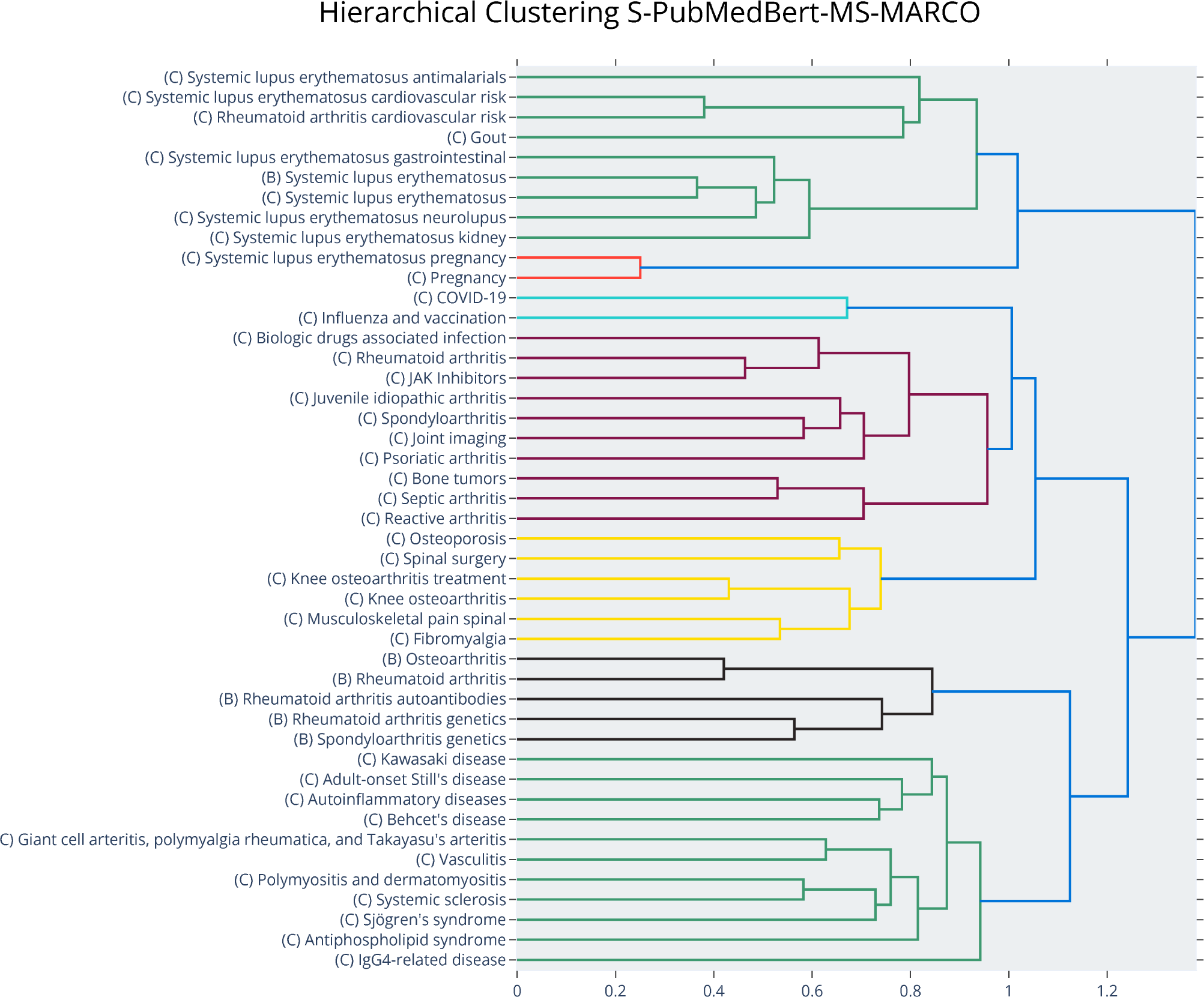
Hierarchical structure of the topics labeled with the agreed label. Best *S-PubMedBert-MS-MARCO* model.

**Supplementary Figure 3:**
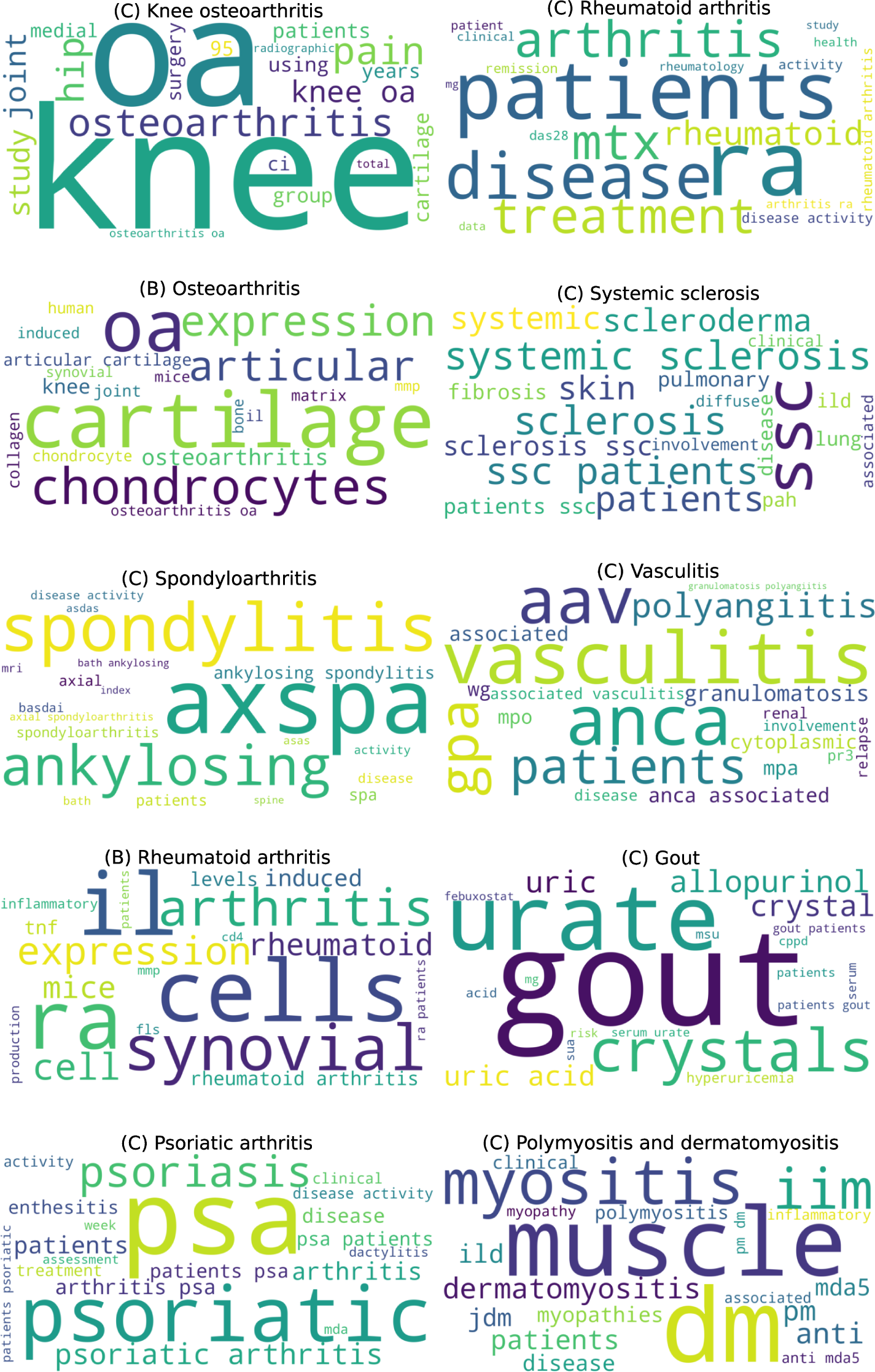
Wordclouds of the top 10 topics of the best *all-mpnet-base-v2* model.

**Supplementary Figure 4:**
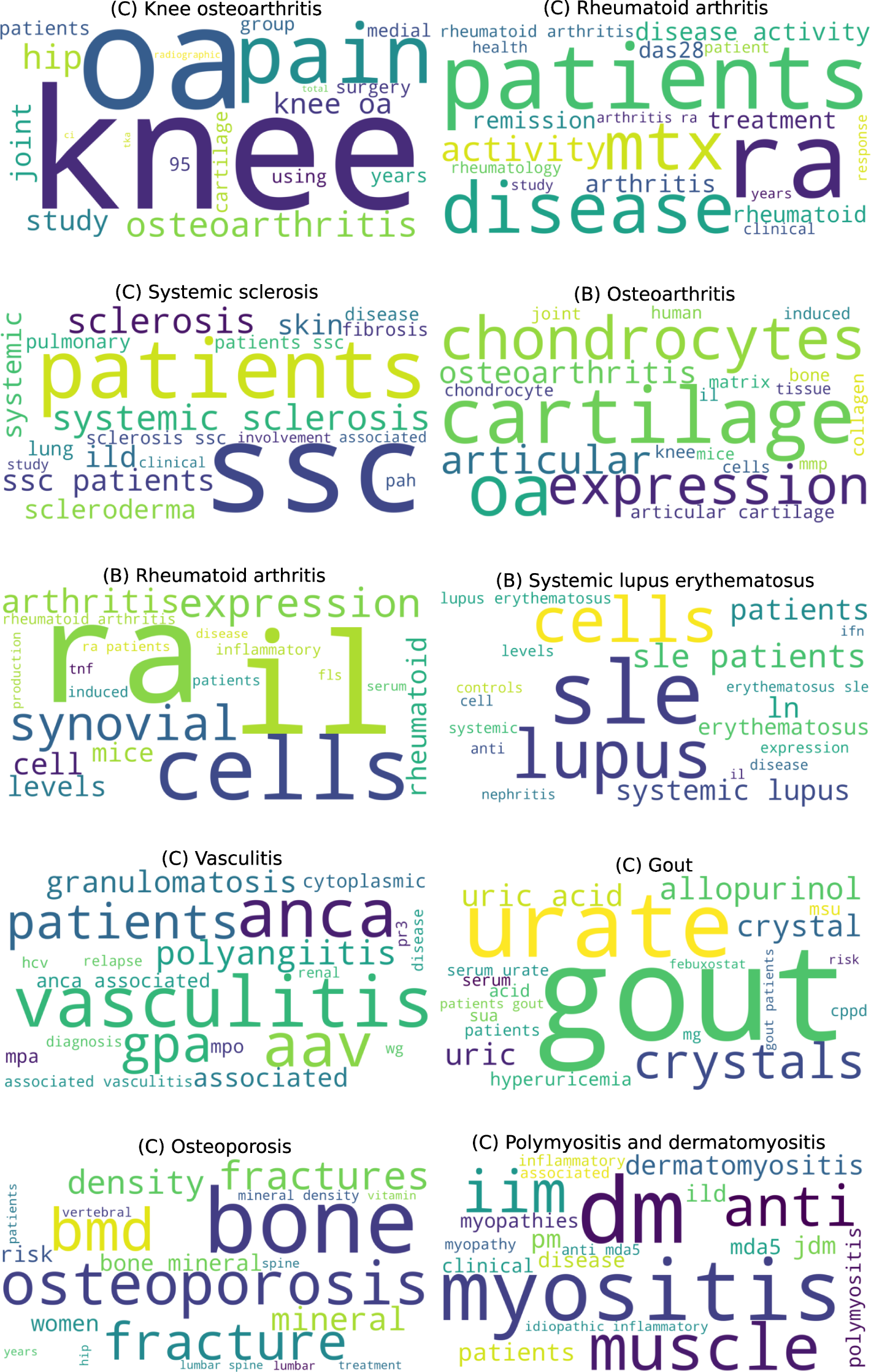
Wordclouds of the top 10 topics of the best *S-PubMedBert-MS-MARCO* model.

**Supplementary Table 1:** Rheumatology journals classified by the Journal Citation Report index as “RHEUMATOLOGY - SCIE”. The jou<u>rnal name is written as appears in JCR webpage. Aktuelle Rheumatologie was exclu</u>ded from this list.

| Journal name |  |
| --- | --- |
| Nature Reviews Rheumatology | Rheumatology and Therapy |
| ANNALS OF THE RHEUMATIC DISEASES | CLINICAL AND EXPERIMENTAL RHEUMATOLOGY |
| Lancet Rheumatology | CLINICAL RHEUMATOLOGY |
| Arthritis & Rheumatology | JCR-JOURNAL OF CLINICAL RHEUMATOLOGY |
| Osteoarthritis and Cartilage | LUPUS |
| RMD Open | Pediatric Rheumatology |
| RHEUMATOLOGY | International Journal of Rheumatic Diseases |
| BEST PRACTICE & RESEARCH IN CLINICAL RHEUMATOLOGY | BMC MUSCULOSKELETAL DISORDERS |
| CURRENT OPINION IN RHEUMATOLOGY | Advances in Rheumatology |
| Current Rheumatology Reports | RHEUMATIC DISEASE CLINICS OF NORTH AMERICA |
| SEMINARS IN ARTHRITIS AND RHEUMATISM | Modern Rheumatology |
| ARTHRITIS RESEARCH & THERAPY | SCANDINAVIAN JOURNAL OF RHEUMATOLOGY |
| ARTHRITIS CARE & RESEARCH | Archives of Rheumatology |
| JOINT BONE SPINE | ZEITSCHRIFT FUR RHEUMATOLOGIE |
| Therapeutic Advances in Musculoskeletal Disease | Acta Reumatologica Portuguesa |
| RHEUMATOLOGY INTERNATIONAL | ARP Rheumatology |
| JOURNAL OF RHEUMATOLOGY |  |
| Lupus Science & Medicine |  |

**Supplementary Table 2:**
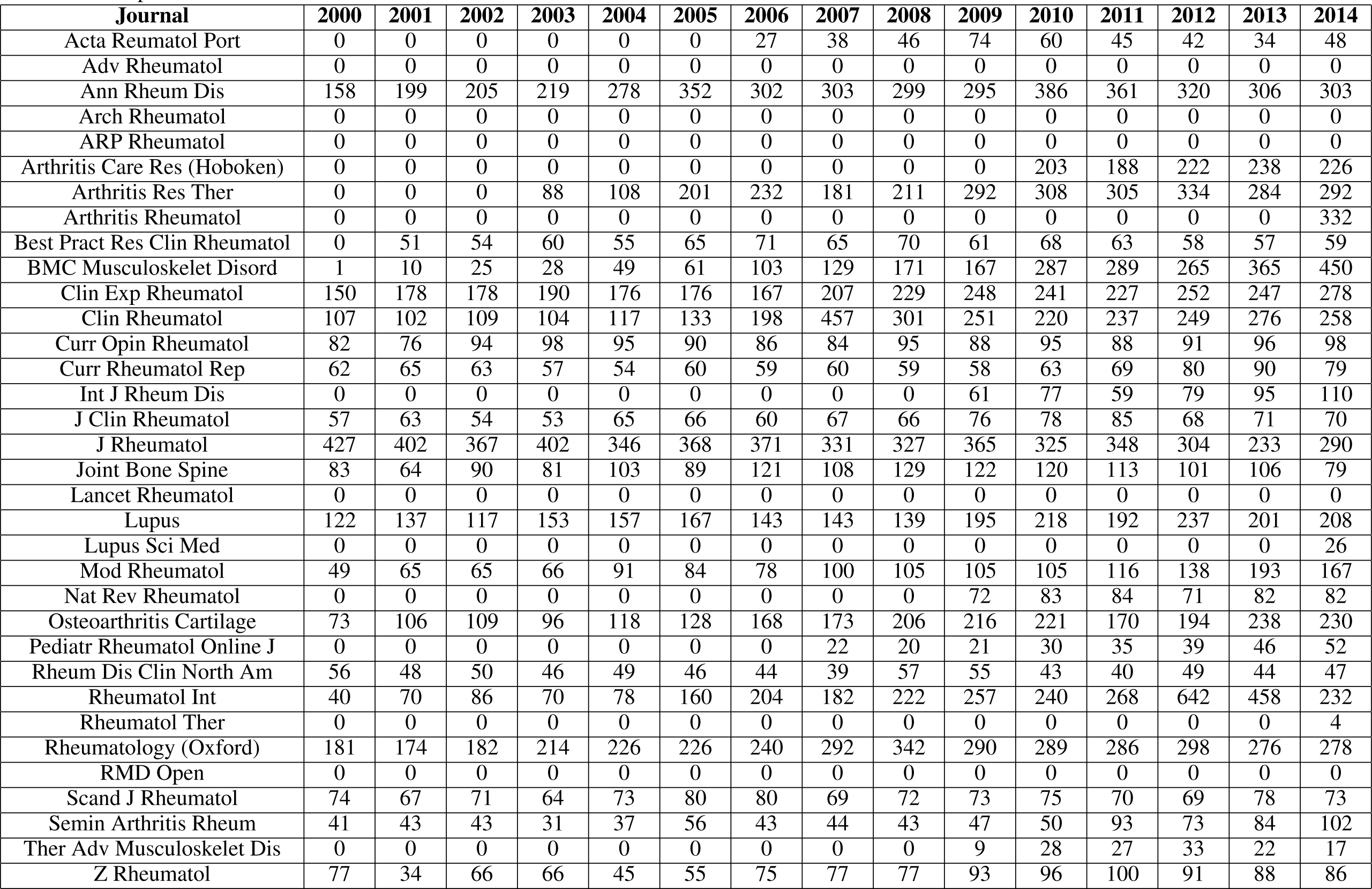
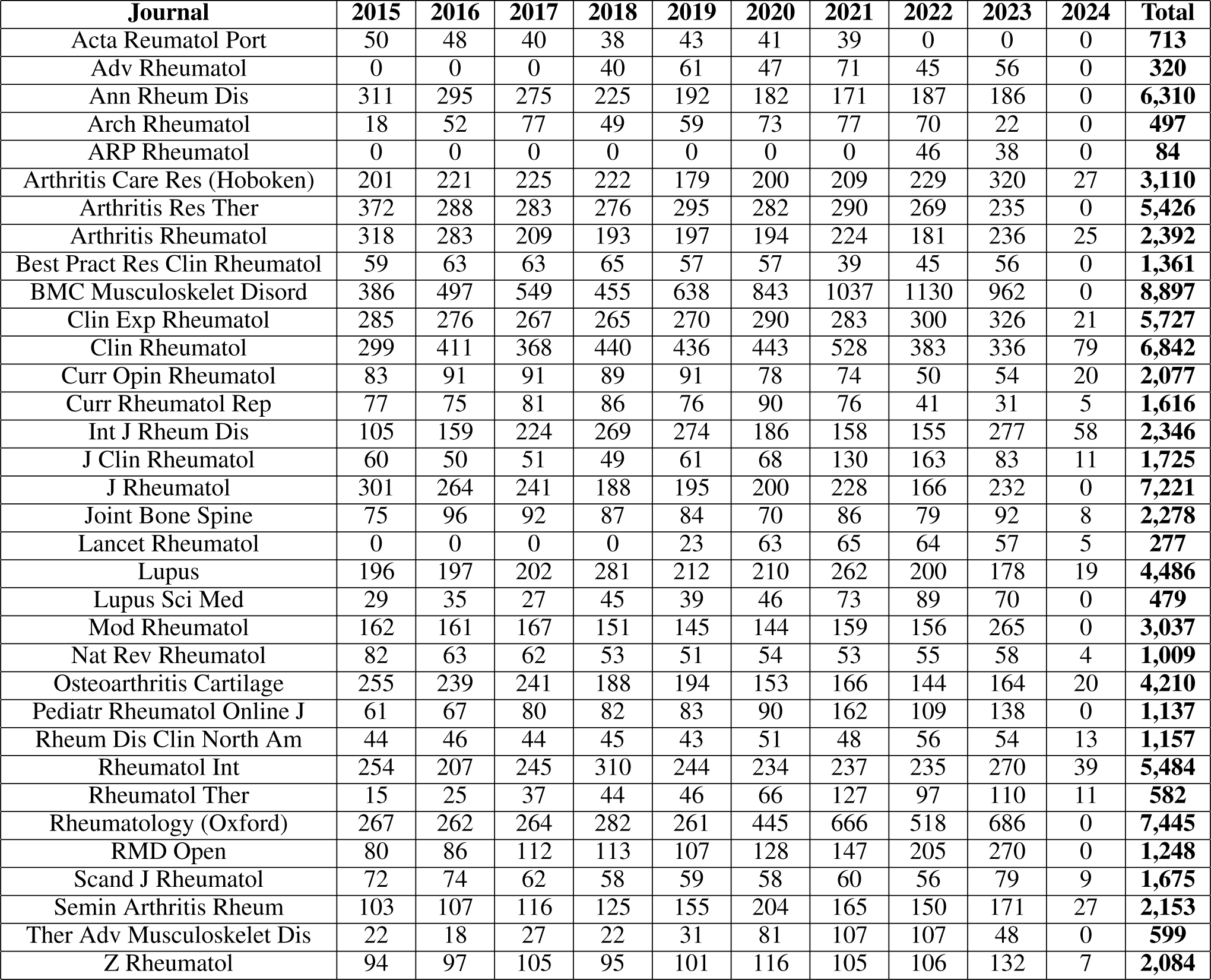
Number of articles with abstract published by year, considering the 34 JCR journals with the category “RHEUMATOLOGY - SCIE”. Although 2024 appears, it must be noted that the time interval studied is 2000-2023. This inconsistency is due to the difference in creation and indexing in PubMed and the date of publication.

**Supplementary Table 3:** Results of the models.

| Model | Min clusters size | Seed | Topics | Outliers | Coherence score |  |  |  |
| --- | --- | --- | --- | --- | --- | --- | --- | --- |
|  |  |  |  |  | u_mass | c_v | c_nmpi | c_uci |
| all-mpnet-base-v2 | 50 | 42 | 296 | 31045 | -0.933 | 0.419 | -0.006 | -2.689 |
| all-mpnet-base-v2 | 50 | 52 | 288 | 30739 | -0.937 | 0.418 | -0.006 | -2.703 |
| all-mpnet-base-v2 | 50 | 62 | 294 | 29168 | -0.923 | 0.419 | -0.006 | -2.713 |
| S-PubMedBert | 50 | 42 | 238 | 33087 | -0.819 | 0.408 | -0.012 | -2.885 |
| S-PubMedBert | 50 | 52 | 246 | 33676 | -0.816 | 0.410 | -0.013 | -2.917 |
| S-PubMedBert | 50 | 62 | 246 | 34309 | -0.823 | 0.409 | -0.011 | -2.875 |
| all-mpnet-base-v2 | 100 | 42 | 151 | 29265 | -0.689 | 0.492 | 0.008 | -2.449 |
| all-mpnet-base-v2 | 100 | 52 | 148 | 30212 | -0.680 | 0.492 | 0.009 | -2.439 |
| all-mpnet-base-v2 | 100 | 62 | 154 | 29951 | -0.689 | 0.489 | 0.009 | -2.454 |
| S-PubMedBert | 100 | 42 | 129 | 35332 | -0.610 | 0.486 | -0.005 | -2.762 |
| S-PubMedBert | 100 | 52 | 126 | 33162 | -0.589 | 0.479 | -0.008 | -2.838 |
| S-PubMedBert | 100 | 62 | 129 | 35102 | -0.606 | 0.492 | 0.002 | -2.601 |
| all-mpnet-base-v2 | 150 | 42 | 87 | 27896 | -0.480 | 0.535 | 0.009 | -2.438 |
| all-mpnet-base-v2 | 150 | 52 | 85 | 26068 | -0.477 | 0.533 | 0.012 | -2.351 |
| all-mpnet-base-v2 | 150 | 62 | 71 | 21245 | -0.417 | 0.521 | -0.003 | -2.692 |
| S-PubMedBert | 150 | 42 | 83 | 35288 | -0.467 | 0.532 | 0.004 | -2.566 |
| S-PubMedBert | 150 | 52 | 84 | 35021 | -0.444 | 0.537 | 0.000 | -2.636 |
| S-PubMedBert | 150 | 62 | 81 | 33608 | -0.461 | 0.537 | 0.005 | -2.537 |
| all-mpnet-base-v2 | 200 | 42 | 63 | 26786 | -0.385 | 0.549 | 0.006 | -2.506 |
| all-mpnet-base-v2 | 200 | 52 | 55 | 21398 | -0.347 | 0.541 | -0.002 | -2.680 |
| all-mpnet-base-v2 | 200 | 62 | 61 | 25178 | -0.361 | 0.547 | 0.005 | -2.548 |
| S-PubMedBert | 200 | 42 | 49 | 23908 | -0.320 | 0.574 | 0.015 | -2.241 |
| S-PubMedBert | 200 | 52 | 56 | 26246 | -0.336 | 0.546 | -0.001 | -2.653 |
| S-PubMedBert | 200 | 62 | 54 | 27647 | -0.330 | 0.562 | 0.004 | -2.537 |
| all-mpnet-base-v2 | 250 | 42 | 44 | 22334 | -0.291 | 0.556 | 0.001 | -2.621 |
| **all-mpnet-base-v2** | 250 | 52 | 48 | 22628 | **-0.279** | 0.561 | -0.002 | -2.664 |
| all-mpnet-base-v2 | 250 | 62 | 42 | 19075 | -0.281 | 0.565 | 0.004 | -2.472 |
| **S-PubMedBert** | 250 | 42 | 46 | 26688 | **-0.288** | 0.558 | -0.004 | -2.686 |
| S-PubMedBert | 250 | 52 | 48 | 28045 | -0.291 | 0.551 | -0.006 | -2.771 |
| S-PubMedBert | 250 | 62 | 48 | 28343 | -0.301 | 0.551 | -0.007 | -2.764 |

**Supplementary Table 4:**
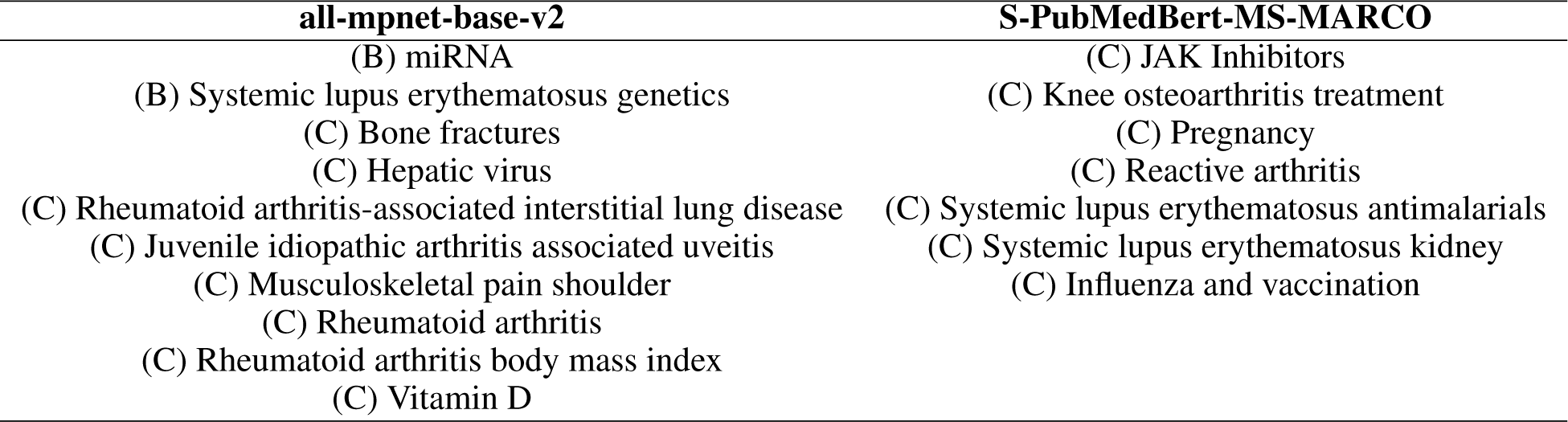
Unique themes of the two selected models.

